# Statistical deconvolution for inference of infection time series

**DOI:** 10.1101/2020.10.16.20212753

**Authors:** Andrew C. Miller, Lauren Hannah, Joseph Futoma, Nicholas J. Foti, Emily B. Fox, Alexander D’Amour, Mark Sandler, Rif A. Saurous, Joseph A. Lewnard

## Abstract

Accurate measurement of daily infection incidence is crucial to epidemic response. However, delays in symptom onset, testing, and reporting obscure the dynamics of transmission, necessitating methods to remove the effects of stochastic delays from observed data. Existing estimators can be sensitive to model misspecification and censored observations; many analysts have instead used methods that exhibit strong bias or do not account for delays. We develop an estimator with a regularization scheme to cope with these sources of noise, which we term the Robust Incidence Deconvolution Estimator (RIDE). We validate RIDE on synthetic data, comparing accuracy and stability to existing approaches. We then use RIDE to study COVID-19 records in the United States, and find evidence that infection estimates from reported cases can be more informative than estimates from mortality data. To implement these methods, we release `incidental`, a ready-to-use `R` implementation of our estimator that can aid ongoing efforts to monitor the COVID-19 pandemic.

---

Information on the progress of an ongoing epidemic arrives with delays. New cases, hospitalizations, and deaths are reported potentially weeks after individuals are infected. This stochastic delay obscures patterns in transmission. Accurate estimation of daily infections is crucial for understanding the dynamics of disease transmission, and assessing the impacts of interventions [11, 14, 29]. However, as we show, currently available methods for reconstructing infection curves exhibit bias and instability.

Mathematically, observations such as daily reported cases can be described as a convolution of the underlying time series of new infections with a delay distribution — the probability distribution that describes time from infection to reporting. Thus, recovering the infections curve from delayed reports is a *deconvolution* operation (Figure 1). Unfortunately, deconvolution of noisy data presents a well-known ill-posed inverse problem, in which signal and noise cannot be separated, even when the delay distribution is known perfectly [24]. Practically, this ill-posedness manifests as instability in estimates, both when the data and assumptions about the delay distribution are perturbed. This instability is compounded in infection estimation by right-censoring — recent infections have a smaller probability of being reported in the observation period. In general, instability in deconvolution problems is addressed by *regularization*, which imposes structure on the signal that is recovered from noisy data [28].

**Figure 1:**
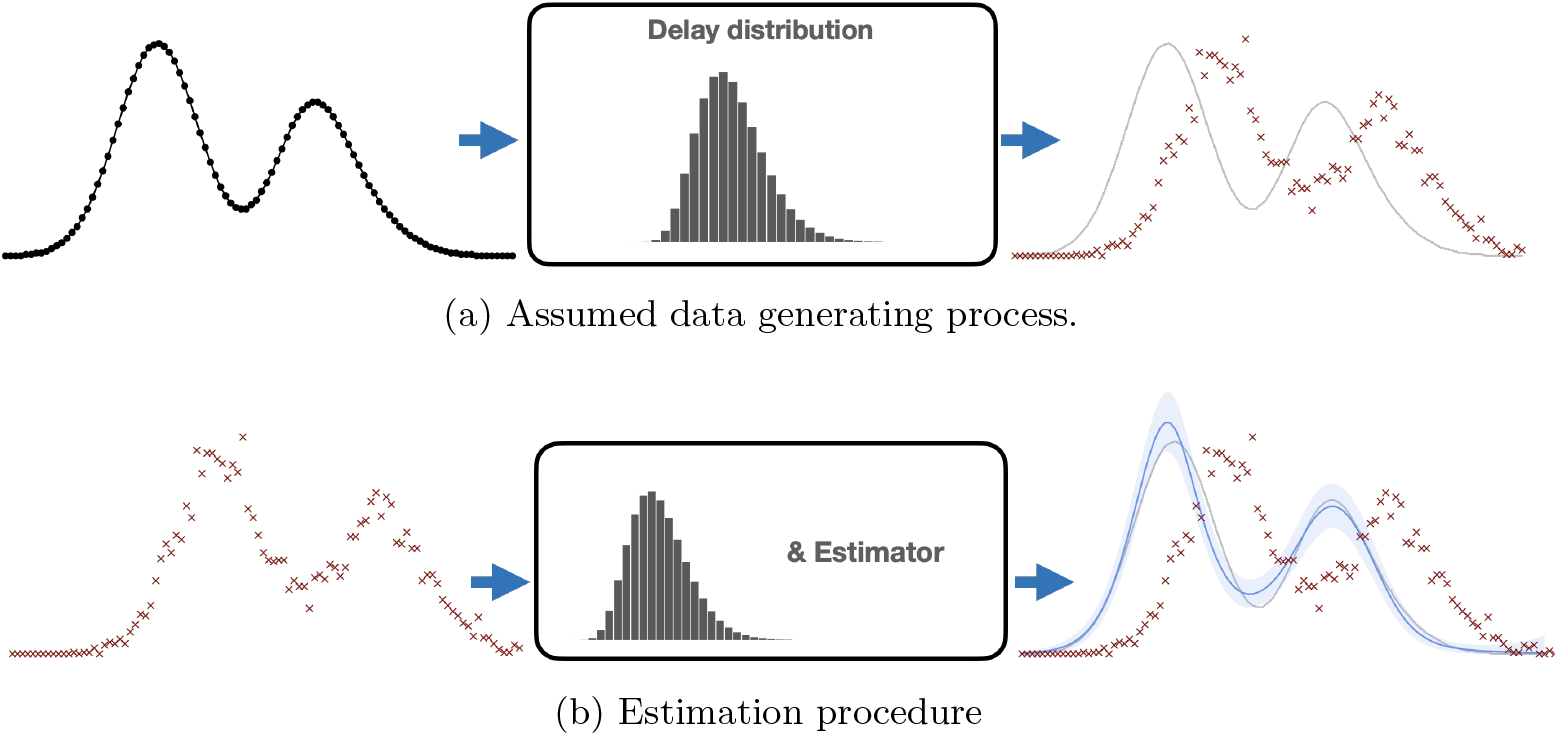
Infection estimation overview. Top: the underlying infection time series — new infections per day — is perturbed by a delay distribution (center) that is measured with other data or assumed known. Each infection date is stochastically delayed, resulting in the reporting curve (red *×* ‘s) — new reported cases per day. Bottom: the estimation procedure aims to undo this stochastic delay. Given observed report curve (left) we use a statistical estimator with the delay distribution to recover the underlying curve.

In this paper, we propose a statistically robust method to infer infection time series from delayed data, which we call the *Robust Incidence Deconvolution Estimator* (RIDE). RIDE incorporates a specific form of regularization that yields stable infection estimates, even in the presence of right censoring. In a simulation study we validate RIDE and compare it to existing methods.

We then use RIDE to study transmission dynamics of SARS-CoV-2 at state and local levels. We compare RIDE to existing estimators on epidemic data from different regions in the United States, qualitatively showing its stability and robustness to censoring. We show that infection estimates from reported cases are often well-aligned with infection estimates from new hospitalizations — a more reliable but less widely available source of epidemic information. Our findings also demonstrate how the implementation and phased lifting of non-pharmaceutical interventions contributed to reductions and increases, respectively, in transmission intensity.

In our simulated and empirical examples, we compare RIDE to two classes of existing methods. The first class, which we term *re-convolution* estimators, estimate the infection curve by sampling from an assumed delay distribution and shifting observed case reports backward in time — effectively, applying a convolution operation in reverse. This heuristic has been applied in a number of public tools for tracking the COVID-19 pandemic [1, 2, 21, 26, 27], but exhibits biases because it is not a deconvolution operation. The second class of estimators perform regularized deconvolution, but yield unstable estimates, in part because their regularization is inadequate in the presence of right-censoring. These include back projection (or back calculation) estimators developed to analyze the AIDS epidemic [7, 8, 9, 10], and the Richardson Lucy (RL) algorithm, a model-free deconvolution method that has been used to analyze influenza [13].

Additionally, we make the proposed method available in an `R` package, `incidental`.

## Results

We first give a brief overview of the statistical estimation problem, existing approaches, and our proposed method, RIDE; we compare these methods in a simulation study. We then study transmission dynamics of SARS-CoV-2, compare infection incidence estimators on real data, and analyze infection time series in relation to non-pharmaceutical interventions.

### Method overview

Given a time series of delayed observations for *T* days, *Y* = (*Y*_1_, …, *Y*_*T*_) (e.g., counts of daily new reported cases) and a delay distribution *θ* = (*θ*_1_, …, *θ*_*P*_) (e.g., the distribution of time from infection to reporting) up to *P* days, the goal is to infer the time series of new infections *X* = (*X*_1_, …, *X*_*T*_). The expected value of the observed data *Y* is a convolution of the infection time series *X* with the delay distribution *θ*; estimation of *X* involves the deconvolution of *Y* and *θ*. To produce an estimator that is robust to noise in *Y*, we propose a model-based estimator using a cubic spline [15] to describe the underlying infections, *X*, and a Poisson likelihood to describe the observed cases. We set the degrees of freedom of the spline basis using AIC [3]. Additionally, we add a regularization penalty on the second difference of the spline parameters, encouraging smoothness and select regularization strength with out-of-sample log likelihood.

Finally, we include an additional regularization to stabilize estimates in the presence of right censoring. Due to delayed reporting, right censoring effectively reduces the number of observations in the most recent time windows, making estimates less stable. We address this issue as a missing data problem, and use a strategy similar to multiple imputation techniques for missing data [22]. Specifically, we sample many extrapolations of the observed time series from a random walk that exhibits the same autocorrelation as the observed data. This random walk encodes the assumption that the autocorrelation in the observed data, which is a direct result of the convolution of infections with the delay distribution, will remain in future observations. For each extrapolation, we condition on the simulated counts to form the incidence estimate, and average estimates across these replicates. We find that this random walk regularization forms much more stable estimates when the report curve is censored. Complete methodological details are given in the *Methods* section and SI A.

### Simulation study

We examine the stability and reconstruction accuracy of estimators on a set of synthetic examples designed to mimic statistical issues associated with COVID-19 data: right censoring and model misspecification. In this simulation study we include re-convolution, Richardson-Lucy, back projection [7, 32] implemented in the `surveillance` package in `R` [17], and our proposed approach RIDE. We compare methods on four synthetic infection curves to study varying levels of complexity: a steep curve with a slow decay, a symmetric curve, a double peaked curve (representing two waves), and a pathological curve that has a sharp climb followed by a total drop-off in cases. For each curve, we consider different observation windows — from highly censored to fully observed — and data simulated under a misspecified delay distribution that approximates processing delays that are common with commercial testing facilities and recording offices. See SI B for a detailed description of synthetic data and results.

In general, we find that the model-based approaches more accurately infer the infection time series than the re-convolution and Richardson-Lucy estimators (as measured by mean squared error). Additionally, we find that RIDE’s regularization scheme is crucial when the model is misspecified; random walk extrapolation is key to stabilizing estimates for highly censored data. Figure 7 summarizes the simulation study.

**Figure 2:**
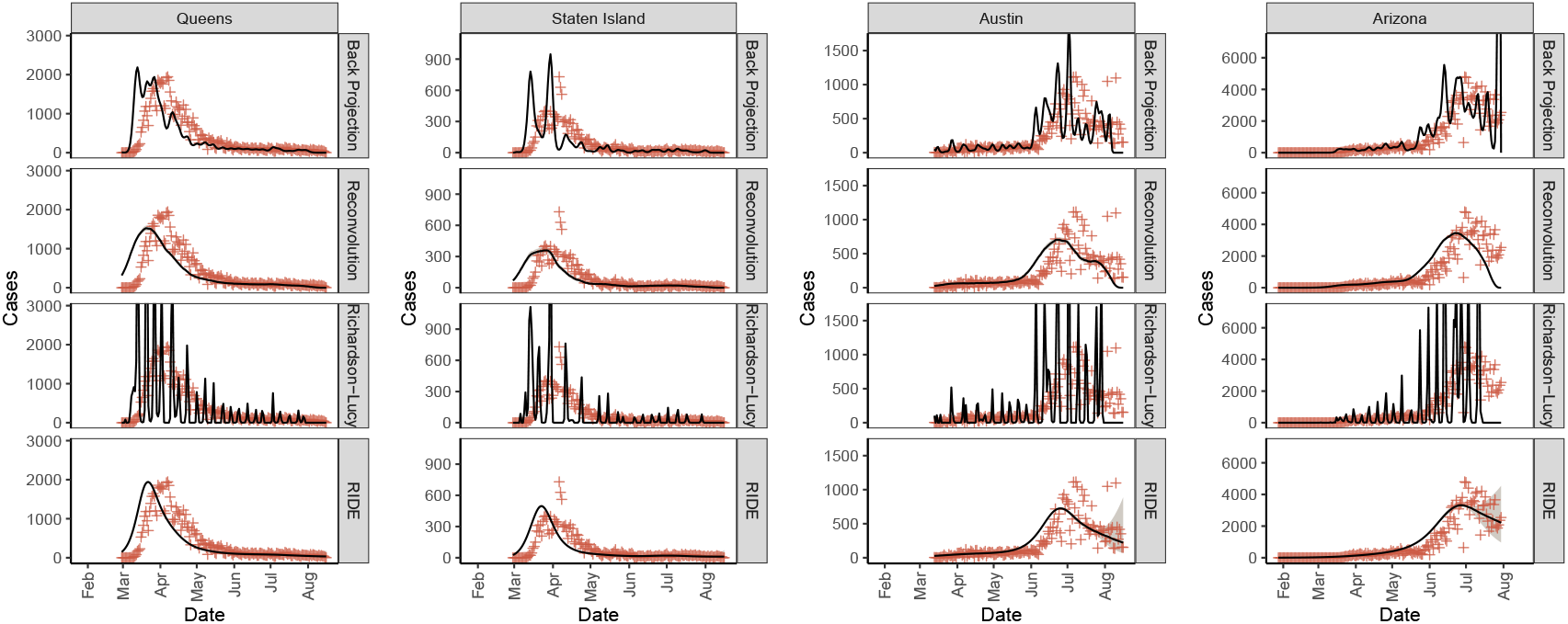
Infections incidence (solid black line), 90% credible regions (gray shaded) when available, and observed values (red plus) by data type across regions (columns) and by method (rows).

**Figure 3:**
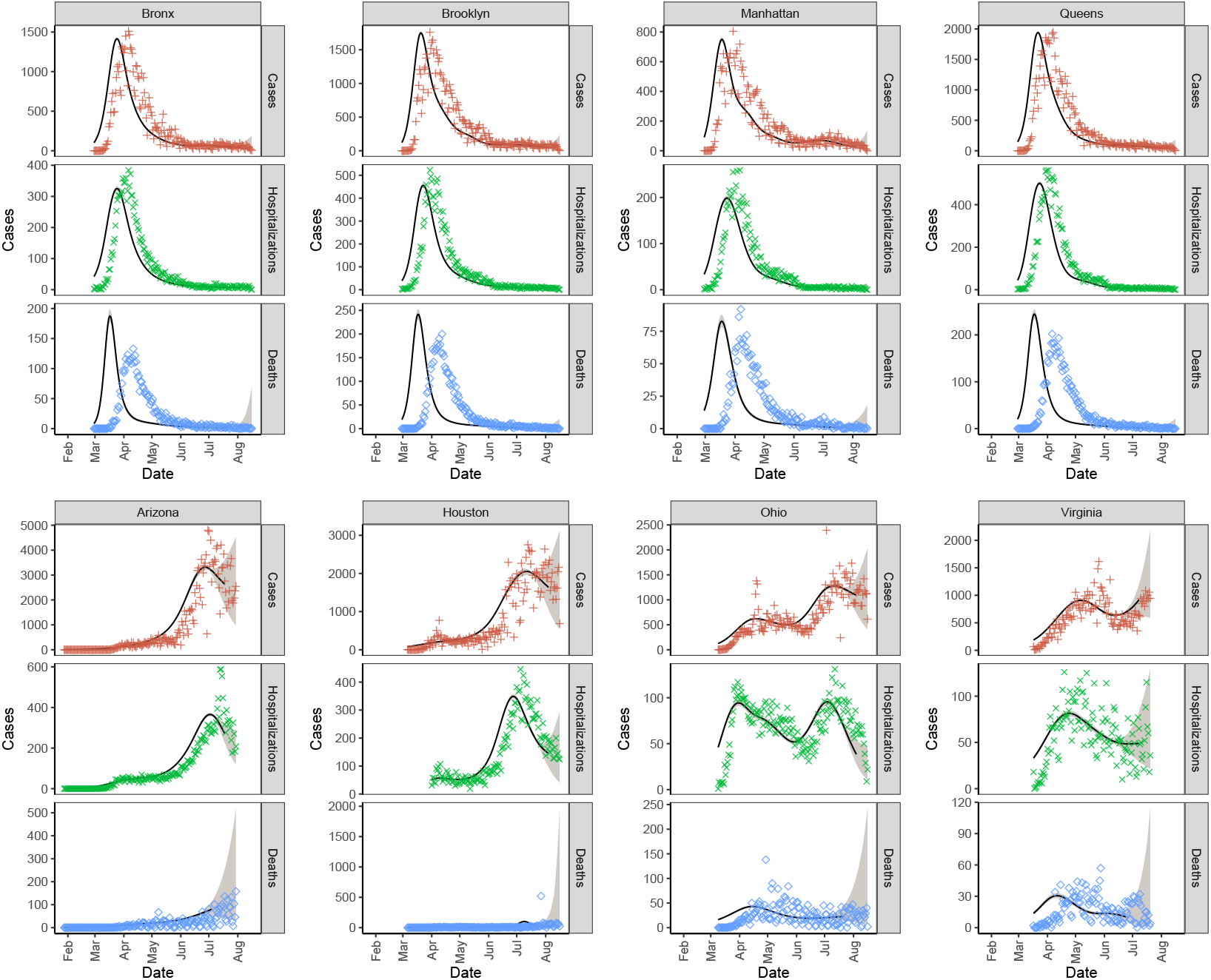
Infections incidence (solid black line), 90% credible regions (gray shaded), and observed values by data type (red plus for cases, green cross for hospitalizations, and blue diamond for deaths) across regions.

**Figure 4:**
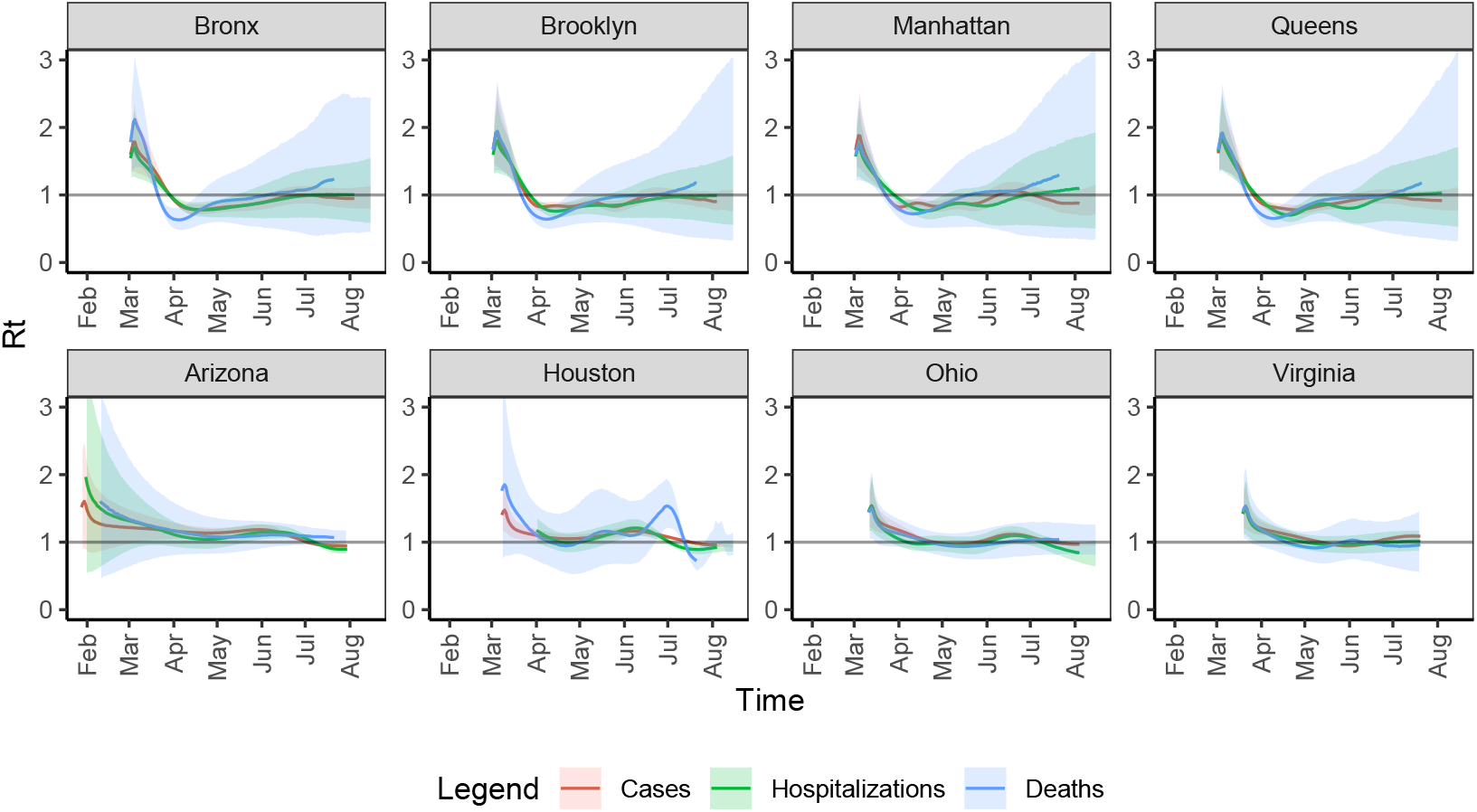
*R*_*t*_ fitted on infection time series estimated by data type across regions. Solid lines are means and ribbons are 90% CIs.

**Figure 5:**
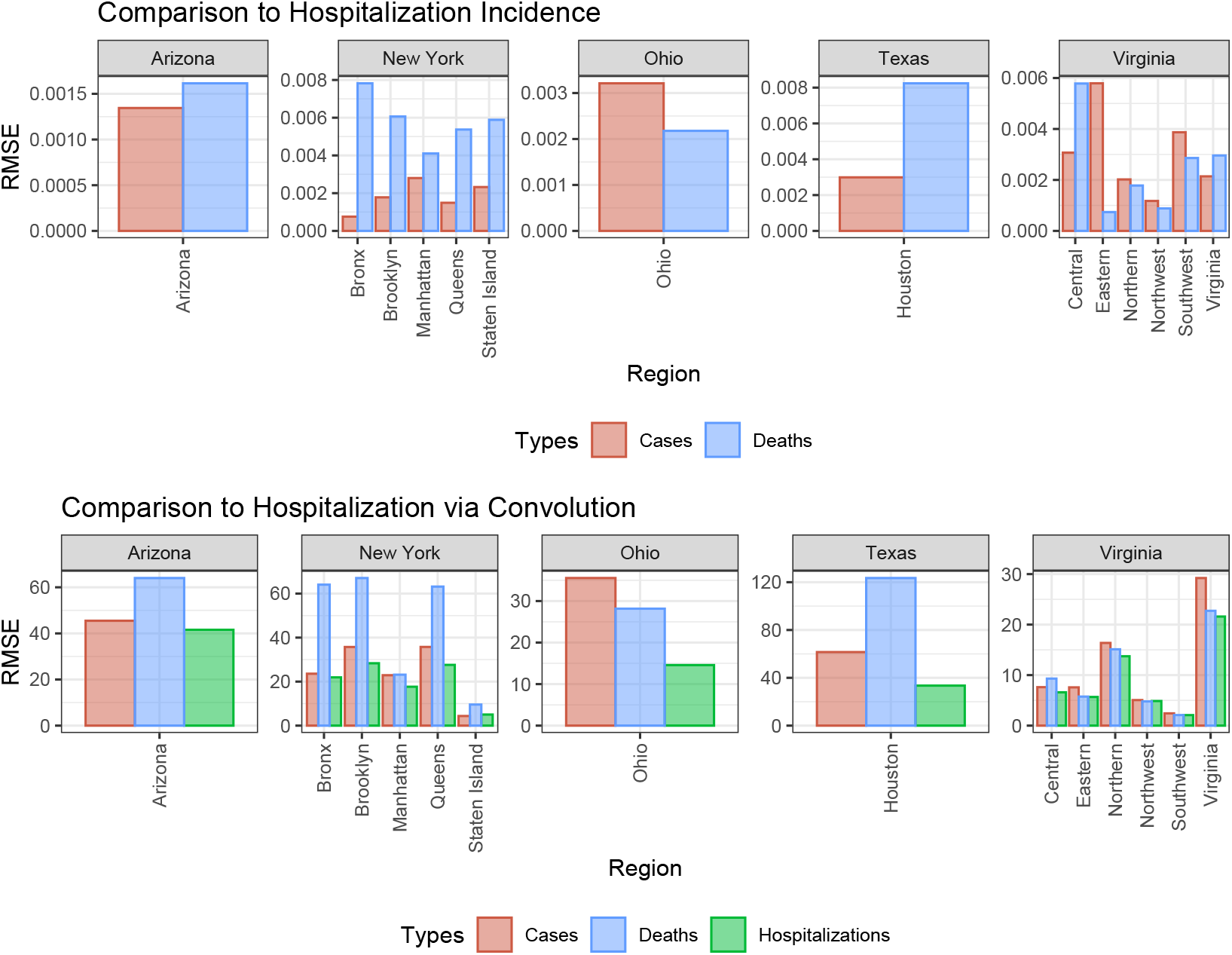
Hospitalization error metrics. The top row is root mean squared error (RMSE) between the normalized hospitalization incidence curve and death/case incidence curves by region. The bottom row is RMSE between the case, hospitalization, and death incidence curves convolved with the hospitalization delay distribution and daily hospitalizations by region.

**Figure 6:**
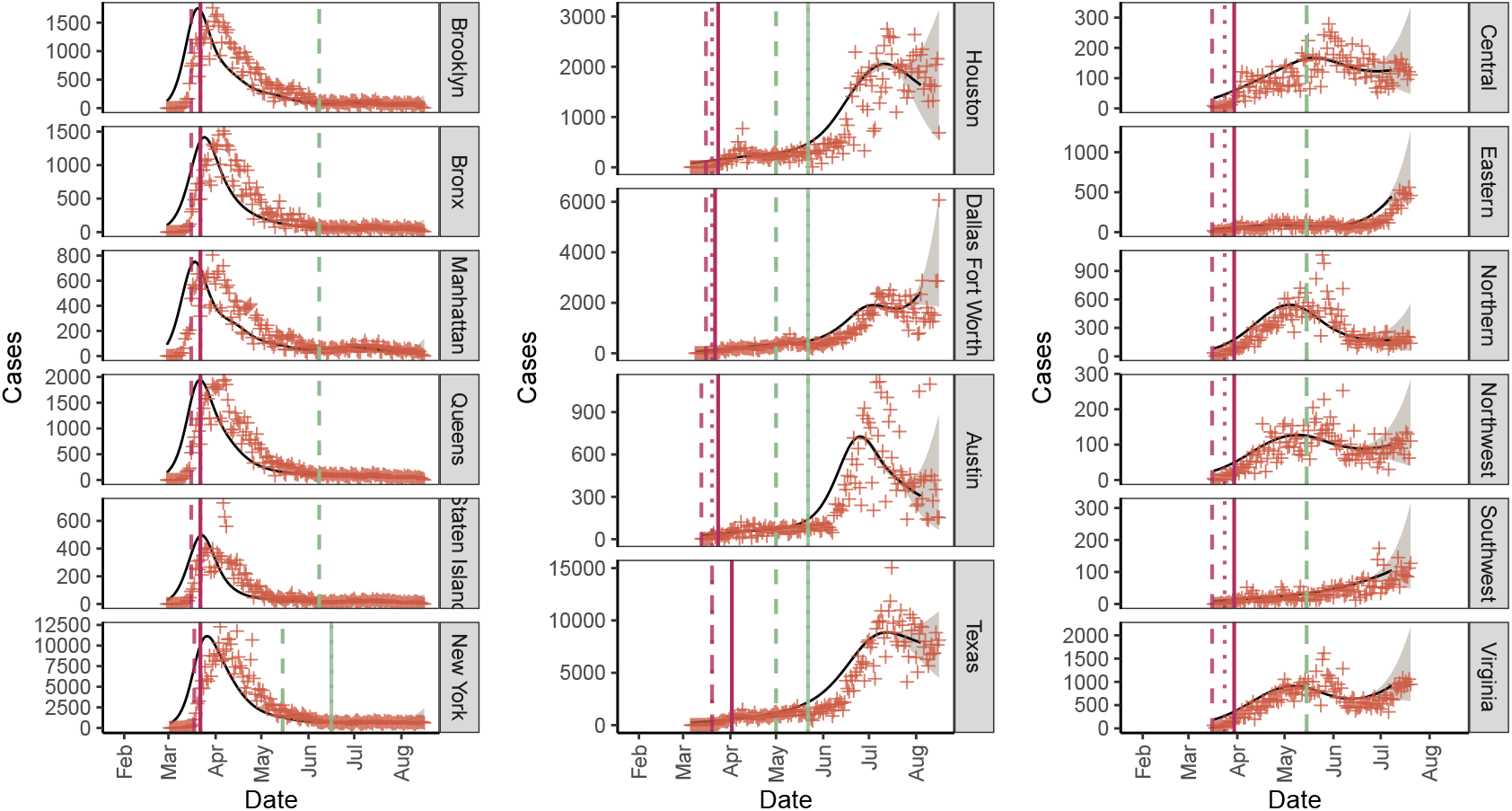
Incidence (solid black line), 90% credible regions (gray shaded), and observed values (red plus) fitted on case data for New York, Texas, and Virginia by sub-region. Policy dates are included: school closures (red thick dash), gatherings ban for 10 or more people (red thin dash), stay-at-home order (red solid line), reopening of low risk non-essential businesses (green thick dash), and bars/restaurants with indoor seating (green solid line).

**Figure 7:**
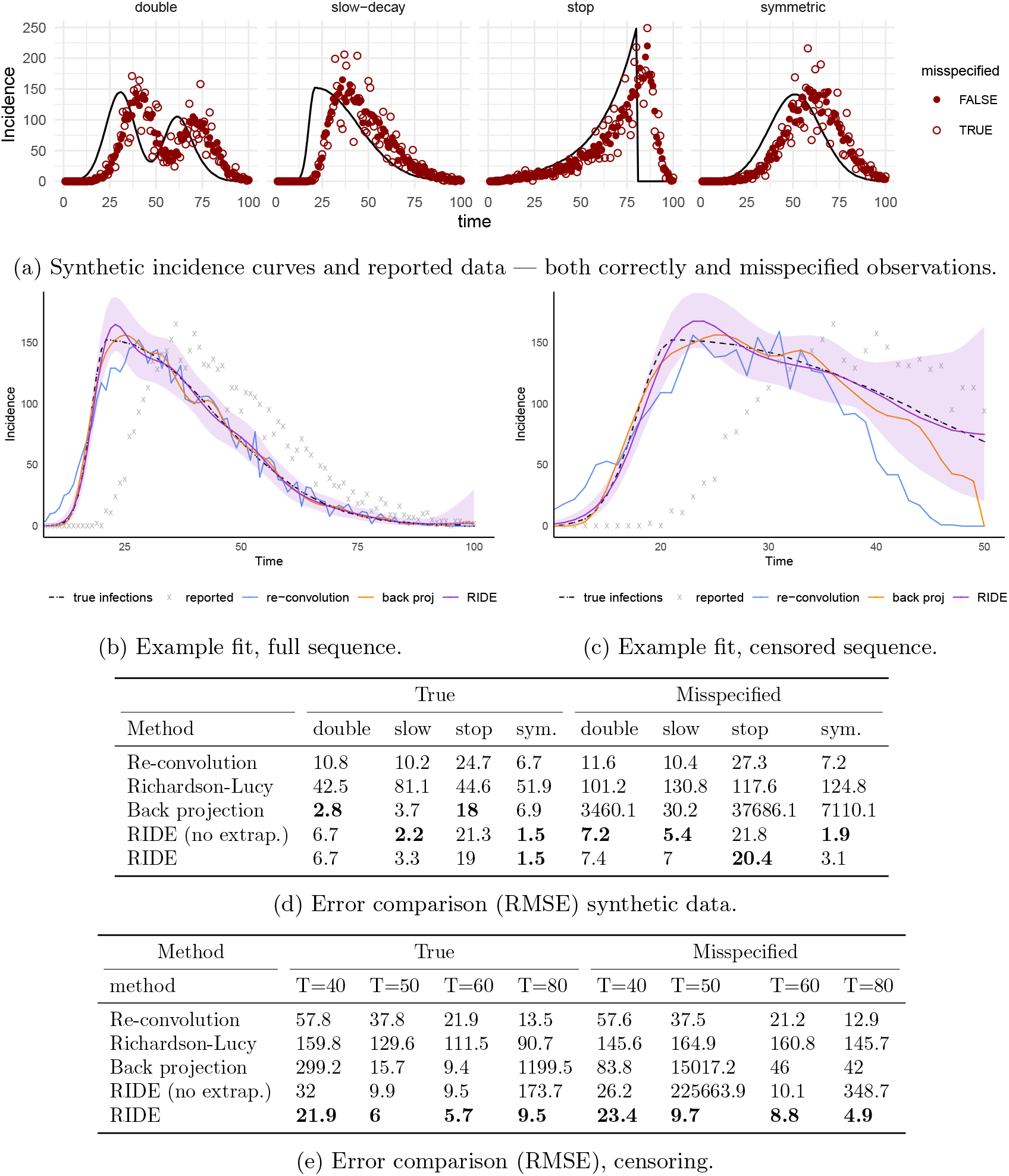
Synthetic experiments. (a) Four synthetic incidence curves and simulated observations with both correct and misspecified noise models. (b) Estimators on the full `slow-decay` data. (c) Estimators on censored `slow-decay` data. (d) Error (RMSE) on all four synthetic curves, for both correct and misspecified data. (e) Error (RMSE) with different levels of censoring on the `slow-decay` synthetic curve. Additional experiment details in SI B and Figures 8-10.

### Application to COVID-19

Monitoring the COVID-19 pandemic presents numerous data challenges [14, 18]. Disease transmission can be studied using different sources of data, each with unique benefits and drawbacks. Data on cases are widely available, with a time delay as short as 12.8 days, on average, from infection to diagnosis [16, 31]; however, such data are greatly affected by widespread variation in testing levels. Data from hospitalizations may be more robust to changes in testing effort [21], but are associated with longer delays, represent a smaller proportion of total cases, and are not widely available in the United States. Deaths suffer the longest delay in both occurrence and reporting, and represent an even smaller proportion of infections. Additionally, improved treatment strategies will likely cause both the infection fatality rate and the duration between infection and death to vary over time.

We aggregated data for daily SARS-CoV-2 tests and new reported COVID-19 cases, hospital admissions, and deaths for four selected regions with high-quality hospital data: Arizona, New York, Ohio, Texas, and Virginia. Hospital admission data is readily available in New York City and Houston within New York and Texas, respectively. In addition to incidence data, we assembled dates for the following policy interventions at the state and regional levels: school closures, bans of gatherings of 10 people or more, stay-at-home orders, reopening of low risk non-essential businesses, reopening of hair salons, and reopening of indoor dining and/or bars. See SI Section C for data sources.

Delay distributions for infection to event are computed by estimating a series of delay times. We estimate time from infection to symptom onset by matching quantiles of a gamma distribution to the data from [19]; from symptom onset to positive test by fitting a gamma distribution to non-zero delay times from Florida for all cases through July 14, 2020^1^; from symptom onset to hospitalization, as fitted by [20] using data from [30, 33]; and from hospitalization to death [20]. See SI Section D for full delay distribution specification.

### Comparison with existing estimation methods

We apply existing estimation methods and the proposed RIDE procedure to COVID-19 case data from Queens, New York, Staten Island, New York, the Austin, Texas metro area, and the state of Arizona in Figure 2. We compare the RIDE procedure output to three existing approaches: back projection [8]^2^, Richardson-Lucy deconvolution [13], and re-convolution [1, 2, 21, 26, 27]. All estimators are described in the *Methods* section in further detail. The re-convolution procedure is a biased estimator and leads to over-smoothing of the infections curve and under estimating the peak. Back projection and Richardson-Lucy are highly sensitive to noise, leading to large oscillations. Further, back projection is sensitive to right censoring — recent infections estimates in Austin and Arizona are highly unstable. The RIDE estimator, on the other hand, is robust to noise in reporting and right censoring.

### Comparison of inferences by data source

We next apply RIDE to case, hospitalization, and mortality data to estimate infections in Arizona, Ohio, Virginia, Houston, and New York City boroughs – locations chosen for their readily available hospitalization data. Figure 3 depicts inferred daily infections (scaled by estimates of the proportion of cases resulting in confirmation, hospitalization, and death, respectively). Note that the peak in estimated infections precedes the peak in reported cases, and has steeper upward and downward trajectories than the observed data time series. Additionally, uncertainty regions near the end of the time series are much larger for daily infections inferred from deaths data. This is due to the longer lag between infection and death relative to cases and hospitalizations. Reproductive numbers fitted using the method of Cori and colleagues [11], based on the inferred infection time series, are given in Figure 4.

Inferred infections from deaths consistently peaked earlier by about a week than inferred infections from hospitalizations in New York City. This may be due to nascent treatment regimes and overwhelmed hospitals in New York City during March and April. We note that *R*_*t*_ estimates based on hospitalization and death inferences track closely in most areas.

Infections inferred from raw case data early in the epidemic (February through April) is often misaligned with hospitalization inferred infections due to low testing levels. There are some changes in testing levels from May onward, but changes in positive rates swamp changes in testing levels for many areas. For some areas, case upswings in June and July 2020 produce lower levels of hospitalization than earlier in the pandemic, likely due to the infected age skewing younger in later months. Reproductive number estimates computed based on infections inferred from hospitalizations and cases are quite close in all areas examined.

We quantitatively evaluate infection estimates from mortality and case data by measuring alignment with infection estimates from hospitalization data. We use two metrics: (i) the distance between the inferred infections curve using hospitalizations and the inferred infections from cases and deaths, and (ii) the distance between observed hospitalizations and imputed hospitalizations from the infections curve inferred from cases, deaths, and hospitalizations — the infections time series convolved with the hospitalization delay distribution. The first metric is a direct comparison of the infections time series from hospitalizations to infections inferred from others sources. The second metric measures a baseline accuracy of the inferred incidence, where no estimate is expected to have lower error than the curve derived from hospitalizations data.

Results are given in Figure 5. Case infections generally, but not always, have lower error in the two metrics. Case infections has much lower error than death incidence in New York, while deaths infections is a better representation of hospitalization incidence in Ohio. This may be due to a cluster of about 4,000 cases related to Ohio prisons in mid-April, which did not result in a corresponding spike in deaths or hospitalizations. When there are changes in testing regimes or population demographics of the infected, inferred curves from differing data sources may diverge. See SI Section E for further details.

### Estimating infections at state and local scales

COVID-19 data — cases, deaths, and hospitalizations — are often reported at the state or national level, which obscures local variation in prevalence and response to policy interventions. We compare inferred infections from cases in the five New York City boroughs with New York state; the Austin, Dallas/Fort Worth, and Houston metros with the state of Texas; and Virginia health regions (Central, Eastern, Northern, Northwest, and Southwest) to statewide across Virginia. Inferred infections are shown in Figure 6. For each region and state, we also include the following policy decisions: school closures, gathering bans for 10 people or more, stay-at-home orders, reopening of low risk non-essential businesses, and reopening of bars or restaurants with indoor seating.

Given the length of the time series fitted and the noise of the case data, slope changes in infections are accurate to within a few days due to estimator smoothing. Our estimator strongly suggests an association between indoor bar/restaurant openings and an increase in transmission in Texas and the southwestern and eastern regions of Virginia. Additionally, we find that in New York City, estimates suggest that the infections incidence decline postdated school closures, but was aligned with statewide stay-at-home orders. Likewise, statewide — but not county level — stay-at-home orders coincided with a local incidence decline in Houston in early April. Earlier stay-at-home orders in Harris county were not aligned with the decline (Figure 6).

## Discussion

COVID-19 transmission dynamics are obscured by the incubation period, testing delays, reporting delays, and time to hospitalization or death. Accurate estimation of the underlying infection time series is a pressing problem rife with challenges and complications, including observation noise, censoring, and model misspecification. Existing methods are either statistically flawed (e.g., re-convolution) or sensitive to noise in reported data (e.g., Richardson-Lucy and back projection). Our proposed estimator, RIDE, is statistically rigorous and robust to some of the data challenges that are present with COVID-19 reported data, including high noise levels and right censoring. Despite concerns that variation in testing effort may make hospitalizations data superior to data from all cases for monitoring infection dynamics, we found that our inferences from case data provided a reasonable proxy for those fit from hospitalization data, which are much less widely available and pose signal-to-noise ratio problems in smaller regions. Case data allow effective estimation of infections at county and regional levels, rather than requiring entire states or countries.

One of the most promising uses for regional infections estimation is as a tool in the evaluation of public policy. Accurate reconstruction of infection time series is necessary to assess how policies influenced transmission over time, in particular when reporting is lagged and multiple interventions may have been undertaken in succession. As we have demonstrated, local COVID-19 dynamics may differ from state-level patterns, and policy decisions are often implemented to mitigate effects in the areas with the highest case loads. Only looking at state-level responses to policy decisions can blur policy effects as areas with different responses are aggregated.

There remains room to improve these estimators. One salient aspect of the ongoing COVID-19 pandemic are day-of-week reporting effects — it is common to see a smaller volume reported on Saturday, Sunday, and Monday in many locations, likely due to increased processing time. This source of error can be incorporated into the likelihood model with additional parameters, introducing a new regularization problem that warrants further study. Additionally, reporting delays can vary by region and change over time. One potential approach for coping with such variation is to jointly model case and hospitalization data and use the relative stability of hospitalization delays to identify changes in the case reporting delay distribution. A joint approach has the potential to more efficiently use all available information, but would be limited to regions where hospitalization data is relatively available. Lastly, it is worth noting that the delay distribution is the single-most important hyperparameter for estimating the infection time series [6]. This delay may change due to reporting habits, improvements in care, or shifting demographics of the infected population, among other factors [4]. Improving estimates hinges on better characterization of these non-stationary effects.

Our results suggest that the method chosen to estimate infection counts influences the estimate itself and conclusions drawn. Providing a stable, accurate, and consistent way to estimate infection time series can enable more accurate characterization of real-time transmissibility (i.e., the effective reproductive number) and ultimately may help policy-makers assess the effectiveness of public health interventions at the state and local levels.

## Methods

We consider the following observation model of individual infected and reported dates. Each individual *n* ∈ {1, …, *N*} who becomes infected on day *I*_*n*_ is confirmed on day *C*_*n*_ ∈ {1, …, *T*}, and we assume that there were no infections prior to the initial time, denoted *t* = 1. The date of confirmation is stochastically delayed from the date of infection *I*_*n*_ ∈ {1, …, *T*},

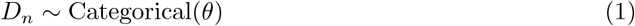

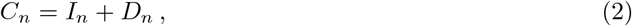

where *D*_*n*_ is a random number of days sampled from a discrete distribution with probability vector *θ* = (*θ*_1_, …, *θ*_*P*_). The infection curve is a time series of daily infection counts, denoted *X* = (*X*_1_, …, *X*_*T*_) where 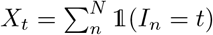 (and 𝟙is the indicator function). Similarly, the observed data is a time series of daily reported cases, denoted *Y* = (*Y*_1_, …, *Y*_*T*_), defined analogously using *C*_*n*_. Our goal is to reconstruct the incidence curve *X* from an observed realization of *Y* = ***y*** = (*y*_1_, …, *y*_*T*_).

We study the performance of different estimators that consider *θ* fixed and known. Practically, COVID-19 line lists track the number of days between symptom onset and the reporting of individual cases, which can be used to form an estimate of the distribution *θ* [16, 31]. We note that this model is a simplification — in practice we observe day-of-week effects in reporting and non-stationarity in *θ*. We examine robustness to model misspecification in our simulation study.

The report curve is related to the incidence curve by a discrete convolution — the expected value of *Y* given *X* can be written as the convolution of *X* and the delay distribution *θ*,

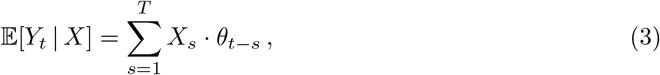

where *θ*_*s*_ = 0 when *s <* 0 or *s > P*. The discrete convolution against *θ* can be expressed as a matrix, *P*_*θ*_, where (*P*_*θ*_)_*t,s*_ ≜ *θ*_*t*−*s*_. The expectation can be written as a matrix multiply 𝔼 [*Y* | *X*] = *P*_*θ*_*X* (see SI Section A for more details). Intuitively, this indicates that to de-convolve the observed signal, an estimator needs to *invert P*_*θ*_.

### “Re-convolution” incidence reconstruction

A popular method for incidence estimation attempts to undo the stochastic delays by sampling from the delay distribution and subtracting the value from each observed time [2, 21, 26, 27]. For each case *n*, the re-convolution estimator samples a delay 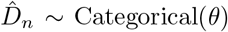 and computes 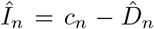, aggregating these into the incidence curve estimate 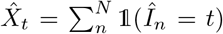. We call this the *re-convolution* estimator, as it amounts to a convolution of the already convolved report curve.

The linear relationship between *X* and *Y* makes clear the conceptual error of the re-convolution method. The re-convolution estimator has the expectation

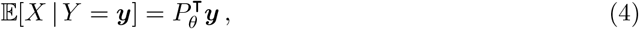

which will be inconsistent in general, as 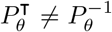. This conceptual error motivates the use of methods developed for *deconvolving* signals (see SI Section A).

### Deconvolution estimators

Deconvolving signals is a well-studied problem in signal processing. One such deconvolution method is the Richardson-Lucy estimator [23, 25], an iterative algorithm. While flexible, this approach can be highly sensitive to observation noise. Nevertheless, the Richardson-Lucy estimator has been used to reconstruct incidence curves for infectious disease [13], and we include the method in our simulation study.

An alternative class of methods uses statistical models to form deconvolved incidence estimates. Back projection (or back calculation) methods are model-based estimators that were developed to infer HIV/AIDS infection incidence [6, 7, 8, 9, 10, 32]. In the HIV/AIDS setting, the incubation period is on the order of months — much longer than the incubation period for seasonal influenza or SARS-CoV-2, necessitating a more statistically rigorous approach. Back projection is closely related to empirical Bayes methods for deconvolution [12]. These approaches form estimates by maximizing the marginal likelihood of observed data given a model for *X*_1_, …, *X*_*T*_ and some form of regularization. Parameterizing the incidence time series as *X*_1_, …, *X*_*T*_ = *X*(*β*) (e.g., with smoothing splines or a step function), a model-based objective function takes the form

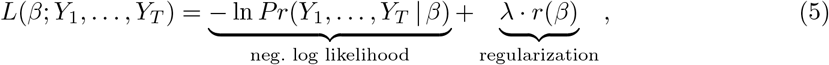

where the likelihood function varies from method to method. Both multinomial [9] and Poisson [7] observation models have been considered; both model-based and post-hoc methods of smoothing have been studied [32].

### Ill-posedness and regularization

Incidence estimation is a deconvolution exercise — a classic ill-posed inverse problem. Specifically, the true incidence curve *X* is *just identified* — the number of free parameters is equal to the number of observed data points [24]. Without observation noise, the convolution matrix *P*_*θ*_ can be inverted, and the true incidence *X* can be identified. With observation noise, however, there may be many plausible incidence curves that explain the observed data. The purpose of regularization is to bias the optimization objective toward estimates with properties we believe to be true, a priori. Smoothness — the belief that incidence should not vary wildly day to day — is the main property induced by regularization.

Here, we devise a model-based estimate using a cubic spline [15] to describe the underlying incidence, *X*(*β*), and a Poisson likelihood to describe the observed cases. Locations with low case counts can be prone to over-fitting due to small sample size. To address this issue, we first select the degrees of freedom of the spline basis using AIC [3]. Additionally, we add an L2 regularization penalty on the second difference of the *β* parameters. To select *λ*, we split the observed data and use out-of-sample log likelihood. By default, we use 25% of the data to estimate out-of-sample log likelihood, and average over four random splits.

The stability of estimates in the most recent time window before *T* is another practical concern. In this window, the effective number of observations is small due to right censoring, and incidence estimates can be unstable without a sensible prior. In this epidemiological setting, the convolution of the incidence and delay distribution results in a time series with significant autocorrelation. We exploit this structure by extrapolating the report curve forward in time with a random walk and condition on these simulated counts to form the incidence estimate. We first apply an Anscombe transform [5] to the observed report curve to stabilize the variance and use the empirical single-lag autocorrelation to simulate random walk extrapolations. We average estimates over replicates of extrapolated random walks in a style similar to common techniques for handling missing data [22]. We find that this random walk regularization forms much more stable estimates when the reported data are censored. We term this procedure the *Robust Incidence Deconvolution Estimator* (RIDE); further details are given in SI Section A.

### Reconstructing known incidence curves from simulated case reports

We study the accuracy of various incidence estimators described herein, including re-convolution, Richardson-Lucy, back projection [7, 32] implemented in the `surveillance` package in `R` [17], and our proposed approach RIDE. The back projection method also employs a Poisson likelihood, but regularizes by injecting a rolling window smoothing step into an expectation maximization fitting procedure. In general, we find that the model-based approaches for incidence estimation are more accurate than the re-convolution and Richardson-Lucy estimators. Additionally, we find that regularization is crucial when the model is misspecified.

We examine four synthetic incidence curves to study varying levels of complexity: a steep incidence curve with a slow decay, a symmetric curve, a double peaked curve (representing two waves), and a pathological curve that has a sharp climb followed by a total drop-off in cases. Additionally, we consider different observation windows from highly censored to fully observed. We assume a delay distribution *Gamma*(*k* = 10, *θ* = 1), with a mean delay of 10 days. Additionally, we study incidence reconstruction for data simulated under a misspecified delay distribution. In the misspecified setting, we mimic weekly reporting delays where on every sixth and seventh day a uniform random proportion of the cases between 30% and 50% are reported two days later. This approximates reporting delays that are common with commercial testing facilities and recording offices. See SI Section B for a detailed description of synthetic data and results. Synthetic data are depicted in Figure 7a.

Inference quality of the methods are evaluated by root mean squared error (RMSE) between the inferred incidence curve and the true incidence curve. Additional metrics are available in the supplement. Performance of each estimator for the four fully observed time series are presented in Figure 7d. The model-based estimators fare better in both the correctly specified and misspecified setting — we observe a consistent improvement in inferred incidence over re-convolution and Richardson-Lucy estimates. Back projection as implemented in the `surveillance` package is competitive in the well-specified setting, but in the misspecified setting accuracy suffers in comparison to our regularization scheme.

We also compare different censoring windows *T* = 40, 50, 60, and 80. Results for the “slow-decay” curve are depicted in Figure 7e. For censored data, random walk regularization stabilizes incidence reconstruction. In the misspecified setting, the Poisson model (and back projection) catastrophically fail for *T* = 50, due to instability in the maximum likelihood solution. The random walk regularization mitigates this pathology while remaining as accurate (or competitive) in all other regimes. We see that both the re-convolution and back projection methods struggle with right censoring. The root of the issue is that neither method accounts for the fact that the reported curve — due to the delayed convolution — is smooth. The incidence curve, therefore, must describe observations we have yet to see. We directly incorporate this into our scheme by sampling potential extrapolations of the report curve, modeled with a simple autoregressive (AR) process. Additionally, averaging over random walk extrapolations yields more realistic uncertainty toward the right of the curve. Figure 8, 9, and 10, illustrate these points in both the correct and misspecified settings.

**Figure 8:**
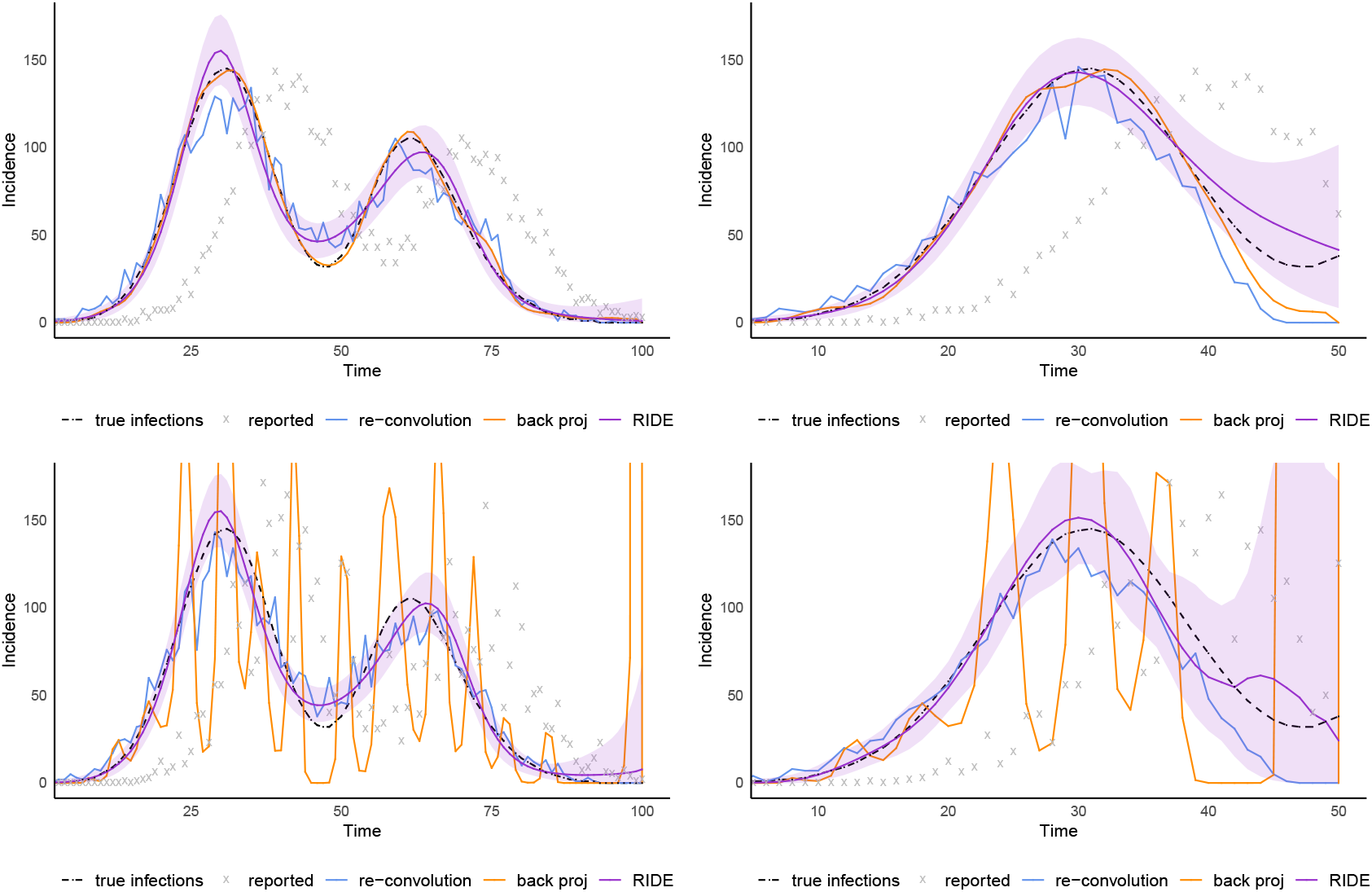
`double`: Top: correctly specified full (left) and censored (right). Bottom: misspecified full (left) and censored (right).

**Figure 9:**
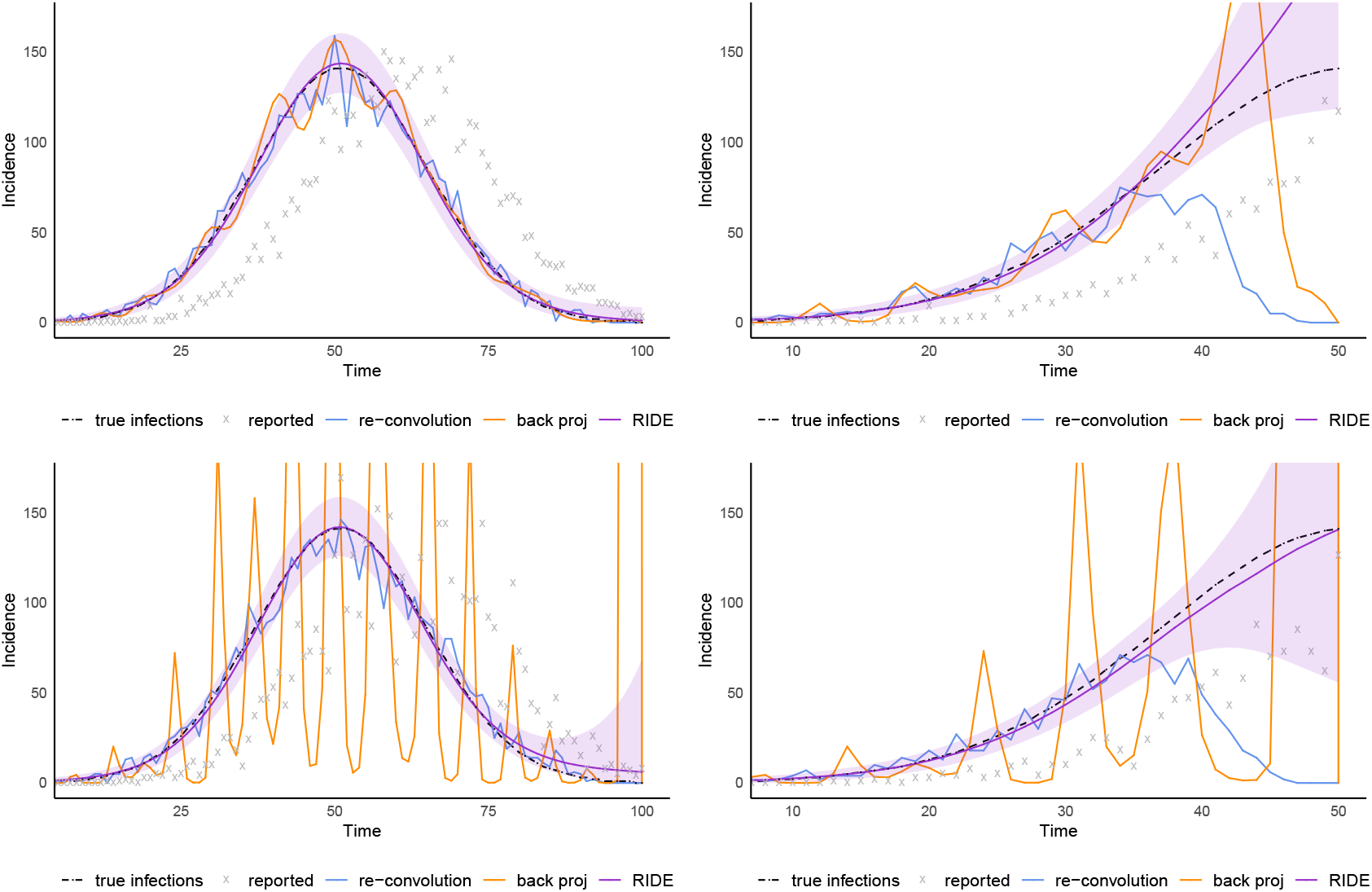
`symmetric`: Top: correctly specified full (left) and censored (right). Bottom: misspecified full (left) and censored (right).

**Figure 10:**
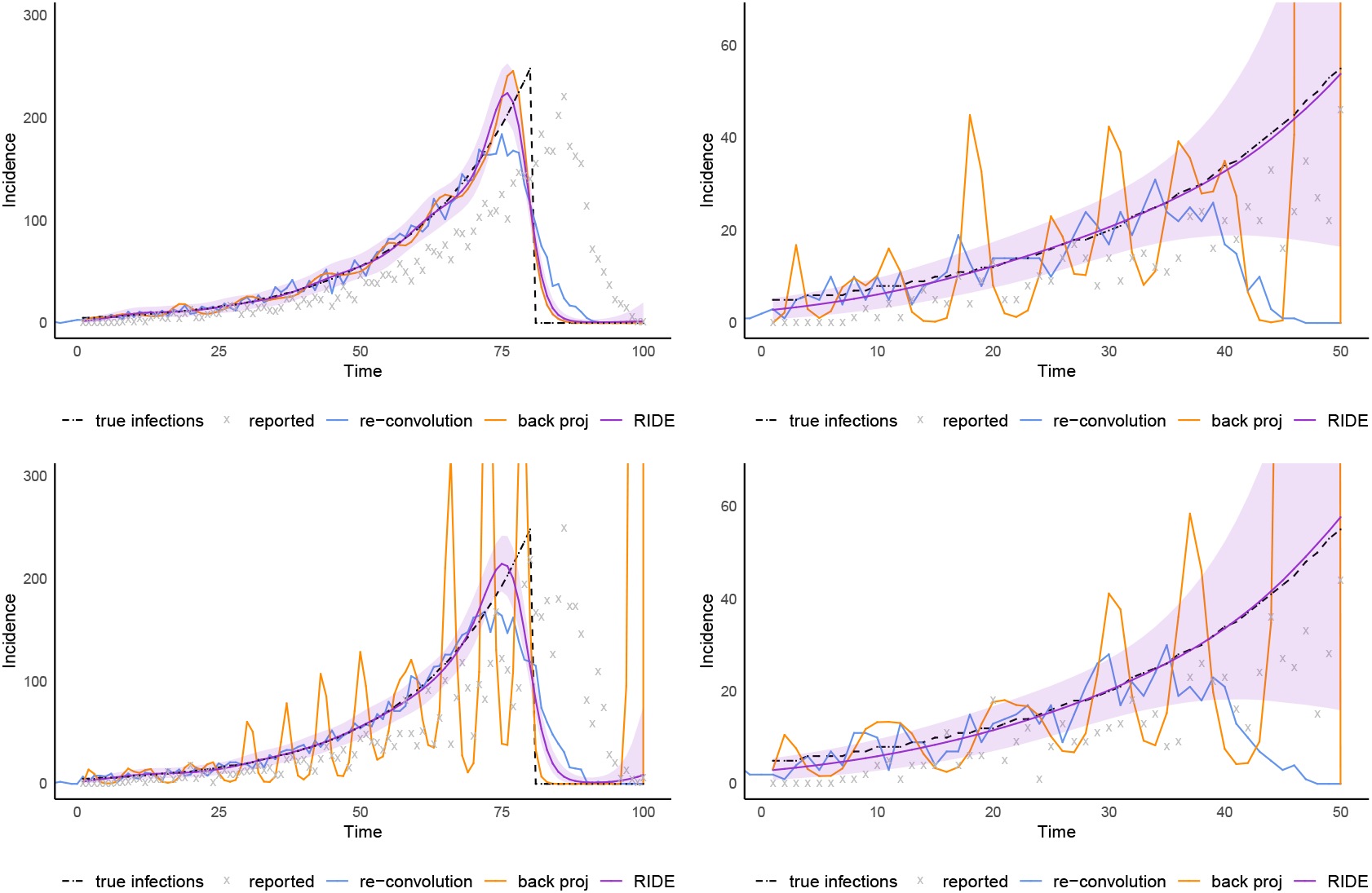
`stop`: Top: correctly specified full (left) and censored (right). Bottom: misspecified full (left) and censored (right).

We use the inferred incidence curves to compute time-varying effective reproductive numbers using the estimator proposed in [11]. Richardson-Lucy deconvolution produces unstable and noisy estimates, leading to reproductive numbers that vary wildly. The re-convolution method over-smooths the incidence curve, leading to systematically biased estimates and typically lower reproductive number estimates in the beginning of the growth cycle. We observe that the model-based estimates more accurately reconstruct the ground truth incidence curve, which translates into more accurate characterization of the time-varying reproductive number.

Additional methodology details are available in SI A.

## Data Availability

Data used in this work are publicly available; sources are identified in the main text and supplemental material.

## Author contribution statement

AD, MS, and RAS identified the problem with reconvolution, proposed deconvolution as an appropriate solution, and noted issues with ill-posedness. AM and LH led the development of the proposed algorithm and its implementation, with input from AD. AM designed and ran the synthetic experiments. LH aggregated publicly available epidemic data, and designed and ran the empirical study. AM led the writing of the manuscript. All authors contributed to designing the research and writing the manuscript.

## A Additional methodology details

We consider the following observation model for infections and confirmed cases. Each individual *n* ∈ {1, …, *N*} who eventually becomes infected is confirmed to be infected on day *C*_*n*_ ∈ {1, …, *T*}, and we assume that there were no infections prior to the initial time, denoted 1. The time of a confirmed case is stochastically delayed from the true date of infection *I*_*n*_ ∈ {1, …, *T*},

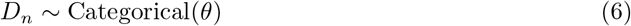

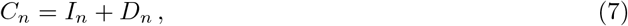

where *D*_*n*_ is a random number of days sampled from a discrete distribution with probability vector *θ* = (*θ*_1_, …, *θ*_*P*_). We assume that *θ* is available as external knowledge from an auxiliary data source and consider it fixed throughout. For example, COVID-19 line lists track the number of days between symptom development and reported case in individual cases can be used to form an estimate of the distribution *θ* [16, 31]. *While the methods considered condition on a specific θ*, uncertainty in *θ* can be explored by a sensitivity analysis or averaging over a distribution of *θ* estimates.

Daily incidence is a summary of all cases from a given day. We write confirmed case incidence on day *t* as 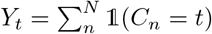, where 𝟙(*C* _*n*_ = *t*) takes the value one when *C* _*n*_ is equal to *t* and zero otherwise. Similarly, the unobserved infection incidence curve is defined 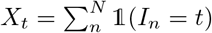. The infection incidence curve is the target of our estimator. We denote realizations of observed daily confirmed case counts as ***c*** = (*c*^(1)^, …, *c*^(*T*)^), and realizations of underlying infection incidence as ***i*** = (*i*^(1)^, …, *i*^(*T*)^). We depict an example of a delay distribution *θ* and the resulting infection incidence and observed incidence pair in Figure 1a.

### Observed case likelihood

The likelihood corresponding to the observed time series, ***y*** is a function of the convolution of the infection incidence *X* = *X*_1_, …, *X*_*T*_ and the delay distribution *θ*. To see this, we view each marginal *Y*_*t*_ as the sum of *N* Poisson processes. Consider the infection date *I*_*n*_ for individual *n*. We can view the value *C*_*n*_ as the realization of a Poisson process with intensity function *θ* offset to start at the value of *I*_*n*_, conditioned on a single realization.

Take all *X*_*s*_ individuals infected on day *s*. Due to the additivity of Poisson processes, the number of cases reported on day *t* that originated on day *s* will follow a Poisson distribution, which we denote

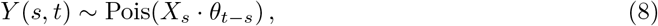

where we define *θ*_*s*_ = 0 for *s <* 0 and *s > P*. The total number of reported cases on day *t* is a sum of *X*(*s, t*) over its first argument up to day *t*, which is again Poisson

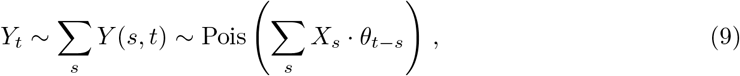

where the mean is the familiar convolution operator. Conditioning on the total number infected, *N*, the vector *C*^(1)^, …, *C*^(*T*)^ is jointly multinomial

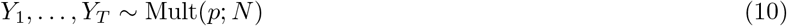

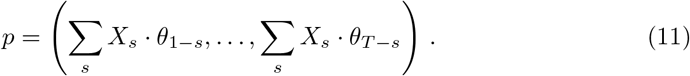

The expected number of reported cases, *E*[*Y*] = (*E*[*Y*_1_], …, *E*[*Y*_*T*_]) is related to the underlying infection time series *X* = (*X*_1_, …, *X*_*T*_) by the convolution

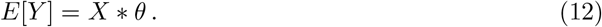

This is equivalent to a linear mapping of *X, E*[*C*] = *P*_*θ*_*X* for the matrix *P*_*θ*_ constructed to correspond to the discrete convolution with vector *θ*

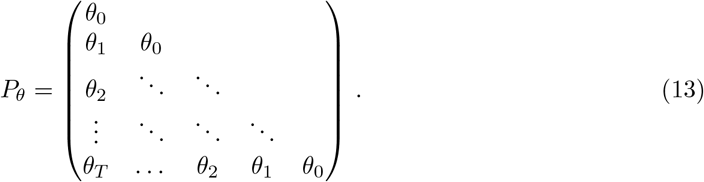

All unidentified entries of *P*_*θ*_ are 0. Intuitively, recovering *X* from *Y* involves the *inverse* of this mapping. However, a popular reconstruction approach does not correspond to inversion.

### “Re-convolution” incidence reconstruction

An intuitive approach to infection incidence reconstruction is to stochastically “undo” the results of the delay random variable, *D*_*n*_. One type of estimator for this is to simply sample a new *D*_*n*_ ∼ Cat(*θ*) from the delay distribution and subtract this new value from the observed case date

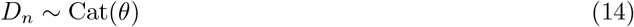

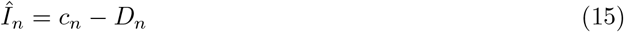

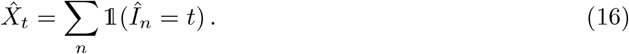

A related approach is to “undo” the convolution of *θ* with *I* by running the convolution backward over the observed case counts, resulting in the estimator

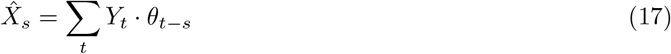

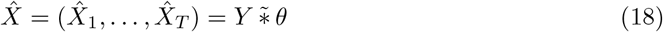

where 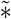 indicates reversing the direction of the convolution operator. Intuitively, this operation distributes observed cases backward in time according to the delay distribution. This has become a popular approach to cope with delayed observations [1, 2, 21, 26, 27].

This reverse convolution corresponds to multiplication by the *transpose* of *P*_*θ*_

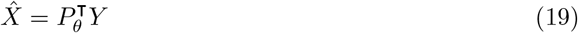

which effectively applies *another* convolution to the already convolved time series *Y*, further smoothing the already smoothed observation.

Given this characterization, a straightforward estimator is to simply invert *P*_*θ*_ to form an estimate of 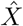.

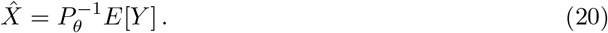

This estimator, however, is not well-defined in the presence of noise in the daily case counts *Y*, rendering it impractical for applied analysis. Intuitively, this inverted convolution estimator is similar to fitting a linear regression model where the number of observations is equal to the number of covariates (i.e., *n* = *p*) — an ill-posed inverse problem [24].

Using the “re-convolution” estimator to impute the incidence curve results in an even smoother imputed curve than the already smoothed reported case incidence curve. Intuitively that makes sense — the “re-convolution” method convolves the already convolved reported case curve with the delay distribution once more, again smoothing (but backward in time).

Using the noiseless (but unobserved) expected reported incidence would recover the underlying daily infections, however applying this estimator to the noisy *observed* reported incidence yields highly unstable estimates — we see the estimate vary between −10^20^ and 10^20^ cases. This instability motivates our development of a practical deconvolution estimator.

### Model-based deconvolution estimators

As stated in the *Methods* section, model-based estimators start with a likelihood model for observed case data, conditioned on the underlying incidence curve. Concretely, model-based methods define an objective

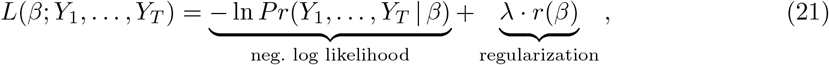

where the form of the log-likelihood is specified by a probabilistic model, and the regularization penalty enforces smoothness in the incidence curve. Back calculation and back projection methods have examined the use of Poisson and multinomial likelihoods [9, 7].

For RIDE, we use a Poisson likelihood and splines for the mean

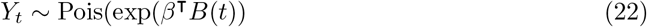

where *β* ∈ ℝ^*P*^ and *B*(*t*) is the *P*-dimensional spline basis value at time *t*.

We also note that the model-based method is closely related to a set of techniques known as empirical Bayes. In fact, the empirical Bayesian method *g*-modeling was developed to de-noise (or deconvolve) noisy observed data [12]. The basic idea behind *g*-modeling is to fit a flexible prior distribution over the unobserved quantity using marginal maximum likelihood. The per-subject unobserved values are the infection times *I*_*n*_ for *n* = 1, …, *N*, and the noisy observations are confirmed case dates *c*_*n*_. The *g*-modeling approach parameterizes the prior (i.e., models the *g*-function), and numerically optimizes this parameter by maximizing the marginal likelihood (i.e., the *f*-function). Specifically we define a prior over *I*_*n*_ parameterized by *β*

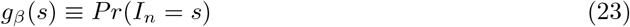

and fit *β* values by maximizing the marginal likelihood of the observed data under the noise model given by *θ*

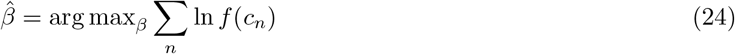

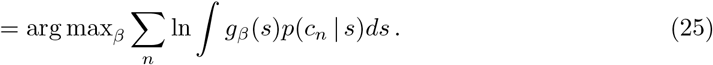

Given this estimate, we can compute the de-noised incidence curve via Bayes rule. As a concrete example, *g*_*β*_(*s*) can be parameterized by a set of basis functions

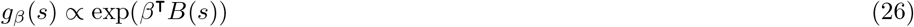

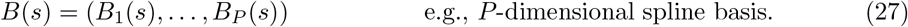

Here, the distribution *g*_*β*_(*s*) is determined by a log-linear function of splines, and is normalized. This empirical Bayes approach leads to a slightly different likelihood — a multinomial. While performance can be similar, we found the Poisson likelihood model easier to fit.

### Regularization

To form estimates, we must choose a set of hyperparameters, including the number of spline basis functions (i.e., the degree of freedom (DoF) parameter, which controls how complex the sample paths can be) and the regularization strength *λ*.

Additionally, to handle censoring, we treat data to the right of the observation window as missing, and impute plausible sample paths, averaging over their uncertainty [22]. Because the smoothing of the stochastic delay typically smooths observations and induces a temporal autocorrelation among reported cases, we extrapolate the report curve with a simple random walk. We first apply an Anscombe transform [5] (i.e., a variance stabilizing transform) and use the empirical single-lag autocorrelation to simulate a set of random walk imputations.

We use a data-driven selection procedure:

- Select spline DoF:
  – Set the default *λ* to be 1*/ Σ* _*t*_ *Y*_*t*_
  – Compute the 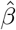 estimate for a grid of DoF
  – Select the DoF parameter with the lowest AIC (or BIC)
- Select *λ* by data splitting — repeat the following four times (by default) and average
  – Split the data into (by default) 25% validation and 75% training by randomly thinning each observed count
  – Compute the 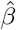 estimate for a grid of candidate *λ* values (with selected DoF value)
  – Select the largest (i.e., smoothest) *λ* within (by default) 2% of the best observed held out log likelihood

The rest of the procedure is as follows: for each imputed time-series, form a set of samples

- Compute the 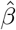 estimate given the selected *λ* and DoF parameters
- Sample *β* from a Laplace approximation formed at the 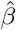 estimate
- Compute the sample path *X*_1:*T*_ for this *β* sample

This procedure yields a collection of *X*_1:*T*_ samples, which we use to form the incidence estimate.

## B Additional Synthetic Data Results

The synthetic models are specified as follows. All of the shapes are normalized to have density 1 and then re-scaled to have 5,000 infections distributed over 100 time steps. All delay distributions are Gamma(10,1). For the well-specified model, each case on the incidence curve is randomly propagated forward *s* days according to the delay distribution. For the noisy (non-well-specified) case, every sixth and seventh day a uniform random number between 0.3 and 0.5 is drawn and that proportion of cases are recorded two days later (e.g. cases from day six move to day eight). This is done to approximate reporting delays for testing and death records.

### Symmetric

Incidence has a Gaussian shape,

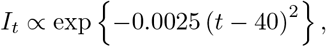

for *t* = 0, …, 89.

### Symmetric Censored

Incidence is the same as Symmetric, except that the incidence curve and reporting stops at *t* = 65.

### Slow Decay

Incidence increases quickly then declines slowly,

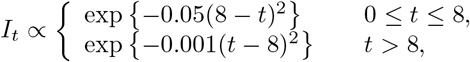

for *t* = 0, …, 88.

### Pathological

Incidence increases quickly then stops entirely,

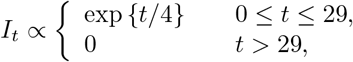

for *t* = 0, …, 79.

### Double Peak

Incidence has two symmetric peaks in succession,

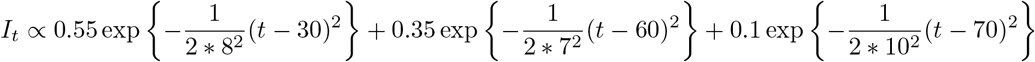

for *t* = 0, …, 100.

## C Data

All case and death data for regions other than New York City is obtained from the New York Times database^3^. Hospitalization data are gathered from other sources in all regions.

### Arizona

Hospitalization data are from the Arizona Department of Health^4^. Test counts and new hospitalization counts are not accurate until 14 days after date reported.

### Ohio

Hospitalization data are from the Ohio Department of Health^5^.

### Texas

Houston hospitalization data is from the Texas Medical Center^6^, which has about 9,200 beds in Houston out of slightly more than 19,000 in total. Areas are defined as follows: Greater Houston area is defined as Austin, Brazoria, Fort Bend, Galveston, Harris, Montgomery, and Waller counties; the Dallas/Fort Worth areas is Collin, Dallas, Denton, Ellis, Hood, Hunt, Johnson, Kaufman, Parker, Rockwall, Somervell, Tarrant, and Wise counties; the Austin area is Travis, Hays, Williamson, Bastrop, and Caldwell counties.

### New York

All New York City data, including cases, deaths, and hospitalizations, is obtained from the New York City Department of Health^7^. State level case and death data is from the New York Times.

### Virginia

Hospitalization data are from the Virginia Department of Health^8^. Regions are state health regions as defined by that department.

## D Delay Distributions

Symptom onset is taken as a proxy for testing date. The testing delay distribution is fit as a Gamma distribution with parameters shape = 4.309 and rate = 0.4668. The hospitalization delay distribution is the symptom delay distribution plus a delay distribution between symptoms and hospitalization, which [20] modeled as a Gamma distribution with shape = 5.078 and rate = 1.307. Finally, we fit a Gamma distribution to the time from hospital admission until death for the distributions presented in [20] using moment matching.

## E Hospitalization Validation

We compute two methods:

1. distance between case, death incidence curves with the hospitalization incidence curve, and
2. distance between observed hospitalizations and the inferred incidence curve from case, death, and hospitalization data convolved with the hospitalization delay distribution.

In both methods, we:

- subset all data to 14 days after hospitalization records begin and 14 days before they end to eliminate ramp up/ramp down effects, and then
- normalize all incidence curves to have a sum of 1 within those ranges.

In the first method, we compute root mean squared error between the normalized hospitalization incidence curve and the normalized method incidence curve,

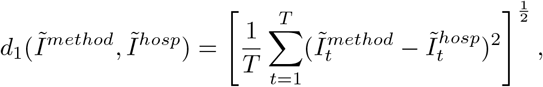

where 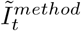 is the normalized incidence curve for a given method on day *t*.

In the second method, we convolve a scaled version of the incidence curve to create an estimate of hospitalizations,

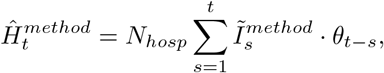

where *N*_*hosp*_ is the total number of hospitalizations. Letting *H*_*t*_ be the observed number of hospitalizations at time *t*,

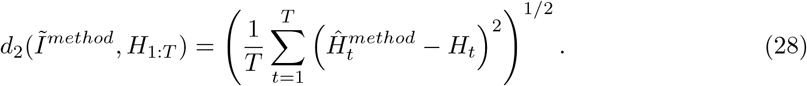

Note that the second method can be used to measure the quality of the incidence curve inferred from hospitalizations. This value should be viewed as a lower bound for other incidence curves.

## Footnotes

1 https://open-fdoh.hub.arcgis.com/datasets/florida-covid19-case-line-data

2 Fit by the backprojNP method in the surveillance package version 1.18.0 in R version 4.0.0 with parameters k=0, eps=rep(0.005,2), iter.max=rep(250,2),B=-1.

3 https://github.com/nytimes/covid-19-data

4 https://www.azdhs.gov/preparedness/epidemiology-disease-control/infectious-disease-epidemiology/covid-19/dashboards/index.php

5 https://coronavirus.ohio.gov/static/COVIDSummaryData.csv

6 https://www.tmc.edu/coronavirus-updates/daily-new-covid-19-cases

7 https://raw.githubusercontent.com/nychealth/coronavirus-data/master/boro/boroughs-case-hosp-death.csv

8 https://www.vdh.virginia.gov/coronavirus/

## References

[1] Sam Abbott, Joel Hellewell, Robin N Thompson, Katharine Sherratt, Hamish P Gibbs, Nikos I Bosse, James D Munday, Sophie Meakin, Emma L Doughty, June Young Chun, et al. Estimating the time-varying reproduction number of sars-cov-2 using national and subnational case counts. Wellcome Open Research, 5(112):112, 2020.

[2] Sam Abbott, Joel Hellewell, Robin N. Thompson, Katherine Sherratt, Hamish P Gibbs, Nikos I Bosse, James D Munday, Sophi Meakin, Emma L Doughty, June Young Chun, Yung-Wai Desmond Chan, Flavio Finger, Paul Campbell, Aira Endo, Carl A B Pearson, Amy Gimma, Tim Russel, CMMID COVID modeling group, Stefan Flasche, Adam J Kucharski, Rosalind M Eggo, and Sebastian Funk. Temporal variation in transmission during the COVID-19 outbreak. https://epiforecasts.io/covid/ (accessed 2020-09-01), 2020.

[3] Hirotugu Akaike. A new look at the statistical model identification. IEEE Transactions on Automatic Control, 19(6):716–723, 1974.

[4] Sheikh Taslim Ali, Lin Wang, Eric HY Lau, Xiao-Ke Xu, Zhanwei Du, Ye Wu, Gabriel M Leung, and Benjamin J Cowling. Serial interval of SARS-CoV-2 was shortened over time by nonpharmaceutical interventions. Science, 369(6507):1106–1109, 2020.

[5] Francis J Anscombe. The transformation of Poisson, binomial and negative-binomial data. Biometrika, 35(3/4):246–254, 1948.

[6] Peter Bacchetti, Mark R Segal, and Nicholas P Jewell. Backcalculation of HIV infection rates. Statistical Science, 8(2):82–101, 1993.

[7] Niels G Becker, Lyndsey F Watson, and John B Carlin. A method of non-parametric back-projection and its application to AIDS data. Statistics in Medicine, 10(10):1527–1542, 1991.

[8] Ron Brookmeyer. Reconstruction and future trends of the AIDS epidemic in the united states. Science, 253(5015):37–42, 1991.

[9] Ron Brookmeyer and Mitchell H Gail. A method for obtaining short-term projections and lower bounds on the size of the AIDS epidemic. Journal of the American Statistical Association, 83(402):301–308, 1988.

[10] Ron Brookmeyer and MitchellH Gail. Minimum size of the acquired immunodeficiency syndrome (AIDS) epidemic in the United States. The Lancet, 328(8519):1320–1322, 1986.

[11] Anne Cori, Neil M Ferguson, Christophe Fraser, and Simon Cauchemez. A new framework and software to estimate time-varying reproduction numbers during epidemics. American Journal of Epidemiology, 178(9):1505–1512, 2013.

[12] Bradley Efron. Empirical Bayes deconvolution estimates. Biometrika, 103(1):1–20, 2016.

[13] Edward Goldstein, Jonathan Dushoff, Junling Ma, Joshua B Plotkin, David JD Earn, and Marc Lipsitch. Reconstructing influenza incidence by deconvolution of daily mortality time series. Proceedings of the National Academy of Sciences, 106(51):21825–21829, 2009.

[14] Katelyn M Gostic, Lauren McGough, Edward Baskerville, Sam Abbott, Keya Joshi, Christine Tedijanto, Rebecca Kahn, Rene Niehus, James A Hay, Pablo M. De Salazar, Joel Hellewell, Sophie Meakin, James Munday, Nikos Bosse, Katharine Sherratt, Robin M Thompson, Laura F White, Jana Huisman, Jérémie Scire, Sebastian Bonhoeffer, Tanj Stadler, Jacco Wallinga, Sebastian Funk, Marc Lipsitch, and Sarah Cobey. Practical considerations for measuring the effective reproductive number, rt. medRxiv, 2020.

[15] Peter J Green and Bernard W Silverman. Nonparametric regression and generalized linear models: a roughness penalty approach. CRC Press, 1993.

[16] Open COVID-19 Data Working Group. Detailed Epidemiological Data from the COVID-19 Outbreak. Accessed on 2020-06-16 from http://virological.org, 2020.

[17] Michael Höhle and Andrea Riebler. The R-package surveillance. 2005.

[18] Nicholas P Jewell, Joseph A Lewnard, and Britta L Jewell. Caution warranted: using the Institute for Health Metrics and Evaluation model for predicting the course of the COVID-19 pandemic. Annals of Internal Medicine.

[19] Stephen A Lauer, Kyra H Grantz, Qifang Bi, Forrest K Jones, Qulu Zheng, Hannah R Meredith, Andrew S Azman, Nicholas G Reich, and Justin Lessler. The incubation period of coronavirus disease 2019 (COVID-19) from publicly reported confirmed cases: estimation and application. Annals of Internal Medicine, 172(9):577–582, 2020.

[20] Joseph A Lewnard, Vincent X Liu, Michael L Jackson, Mark A Schmidt, Britta L Jewell, Jean P Flores, Chris Jentz, Graham R Northrup, Ayesha Mahmud, Arthur L Reingold, et al. Incidence, clinical outcomes, and transmission dynamics of severe coronavirus disease 2019 in California and Washington: prospective cohort study. BMJ, 369, 2020.

[21] Joseph A Lewnard, Vincent X Liu, Michael L Jackson, Mark A Schmidt, Britta L Jewell, Jean P Flores, Chris Jentz, Graham R Northrup, Ayesha Mahmud, Arthur L Reingold, Maya Petersen, Nicholas P Jewell, Scott Young, and Jim Bellows. Incidence, clinical outcomes, and transmission dynamics of hospitalized 2019 coronavirus disease among 9,596,321 individuals residing in California and Washington, United States: a prospective cohort study. medRxiv, 2020.

[22] Roderick JA Little and Donald B Rubin. Statistical analysis with missing data, volume 793. John Wiley & Sons, 2019.

[23] Leon B Lucy. An iterative technique for the rectification of observed distributions. The Astronomical Journal, 79:745–754, 1974.

[24] Finbarr O’Sullivan. A statistical perspective on ill-posed inverse problems. Statistical Science, 1(4):502–518, 1986.

[25] William Hadley Richardson. Bayesian-based iterative method of image restoration. Journal of the Optical Society of America, 62(1):55–59, 1972.

[26] Timothy W Russell, Joel Hellewell, Christopher I Jarvis, Kevin Van Zandvoort, Sam Abbott, Ruwan Ratnayake, Stefan Flasche, Rosalind M Eggo, W John Edmunds, Adam J Kucharski, et al. Estimating the infection and case fatality ratio for coronavirus disease (COVID-19) using age-adjusted data from the outbreak on the Diamond Princess cruise ship, February 2020. Eurosurveillance, 25(12):2000256, 2020.

[27] Kevin Systrom and Thomas Vladeck. R_t_ Covid-19. https://rt.live (accessed 2020-06-16), 2020.

[28] Andrey N. Tikhonov and Vasiliy Y. Arsenin. Solutions of ill-posed problems. Scripta series in mathematics. V.H. Winston and Sons (distributed by Wiley), New York, 1977.

[29] Jacco Wallinga and Peter Teunis. Different epidemic curves for severe acute respiratory syndrome reveal similar impacts of control measures. American Journal of Epidemiology, 160(6):509–516, 2004.

[30] Dawei Wang, Bo Hu, Chang Hu, Fangfang Zhu, Xing Liu, Jing Zhang, Binbin Wang, Hui Xiang, Zhenshun Cheng, Yong Xiong, et al. Clinical characteristics of 138 hospitalized patients with 2019 novel coronavirus–infected pneumonia in Wuhan, China. Journal of the American Medical Association, 323(11):1061–1069, 2020.

[31] Bo Xu, Bernardo Gutierrez, Sumiko Mekaru, Kara Sewalk, Lauren Goodwin, Alyssa Loskill, Emily Cohn, Yulin Hswen, Sarah C. Hill, Maria M Cobo, Alexander Zarebski, Sabrina Li, Chieh-Hsi Wu, Erin Hulland, Julia Morgan, Lin Wang, Katelynn O’Brien, Samuel V. Scarpino, John S. Brownstein, Oliver G. Pybus, David M. Pigott, and Moritz U. G. Kraemer. Epidemiological data from the COVID-19 outbreak, real-time case information. Scientific Data, 7(106), 2020.

[32] Paul SF Yip, KF Lam, Ying Xu, PH Chau, Jing Xu, Wenhu Chang, Yingchun Peng, Zejun Liu, Xueqin Xie, and HY Lau. Reconstruction of the infection curve for SARS epidemic in Beijing, China using a back-projection method. Communications in Statistics—Simulation and Computation, 37(2):425–433, 2008.

[33] Juanjuan Zhang, Maria Litvinova, Wei Wang, Yan Wang, Xiaowei Deng, Xinghui Chen, Mei Li, Wen Zheng, Lan Yi, Xinhua Chen, et al. Evolving epidemiology and transmission dynamics of coronavirus disease 2019 outside Hubei province, China: a descriptive and modelling study. The Lancet Infectious Diseases, 2020.

